# High coverage, persistent gaps: quality of Antenatal Care and its determinants in Zambia based on the 2024 Demographic and Health Survey

**DOI:** 10.64898/2026.06.11.26355447

**Authors:** Paddy Mutungi Tukamuhebwa, Lilian Nuwabaine

## Abstract

**Background:** Evaluating antenatal care (ANC) quality is critical to reducing maternal and neonatal mortality. In Zambia, despite high basic ANC attendance, comprehensive national evidence on the clinical content and quality of services remains limited. This study assessed the coverage of WHO-recommended ANC interventions and identified factors associated with care quality using the latest national data.

**Methods:** A cross-sectional analysis was conducted using data from the 2024 Zambia Demographic and Health Survey. The final analytic sample comprised 4,829 women aged 15–49 with a live birth in the preceding 5 years. A composite index of 15 selected, equally weighted WHO-recommended components evaluated clinical assessment, counseling/screening, preventive interventions, and utilization. Survey-weighted Poisson regression estimated adjusted incidence rate ratios (aIRRs) for the count of ANC components received.

**Results:** The mean ANC quality score was 12.5 out of 15 (95% CI: 12.4–12.6), and 78.5% (95% CI: 77.0–80.0) of women achieved adequate ANC (≥ 12/15 components). While individual clinical and counseling coverage generally exceeded 90%, only 47.2% (95% CI: 45.3–49.0) of women initiated care during the first trimester, and just 4.8% (95% CI: 4.1–5.6) achieved ≥ 8 ANC contacts. Maternal education was the strongest and most stable predictor of quality across all models. Compared to no education, higher education was associated with an 8.0% higher expected quality score (aIRR = 1.080, 95% CI: 1.051–1.110). Lower ANC quality was significantly associated with unwanted pregnancies (aIRR = 0.970, 95% CI: 0.956–0.993) and with residence in Western (aIRR = 0.923, 95% CI: 0.897–0.951) and North Western (aIRR = 0.966, 95% CI: 0.937–0.996) provinces. Absence of distance barriers and residence in Eastern, Luapula, and Copperbelt provinces were associated with higher quality scores.

**Conclusion:** While average ANC component coverage in Zambia is high, critical gaps persist in early initiation and total contact frequency. Care adequacy is strongly influenced by maternal education, relationship status, pregnancy intention, and regional inequities. These findings underscore the need for interventions targeted at uneducated women, preventing unintended pregnancies, and underserved regions such as Western and North Western Provinces.

## Introduction

Sub-Saharan Africa continues to bear the disproportionate global burden of maternal and neonatal mortality, accounting for nearly two-thirds of global maternal deaths [1]. Under the United Nations Sustainable Development Goals (SDG 3), target 3.1 aims to reduce the global maternal mortality ratio to less than 70 deaths per 100,000 live births by 2030, while target 3.2 targets a reduction in neonatal mortality to at least 12 deaths per 1,000 live births [1].

Approximately 210 million pregnancies occur worldwide each year; the associated risks and complications can be significantly mitigated through high-quality antenatal care (ANC) [2]. Regular and comprehensive ANC encounters provide a vital platform for essential healthcare functions, including risk identification, management of pregnancy-related diseases, health promotion, and birth preparedness [1].

Antenatal care (ANC) provides a critical platform for delivering evidence-based interventions to detect and manage pregnancy-related complications, promote healthy behaviours, and link women to essential maternal and newborn health services [3]. In 2016, the World Health Organization (WHO) issued updated recommendations on routine ANC, shifting from a minimum of 4 focused visits to a model of at least 8 contacts and emphasising a comprehensive package of nutritional support, maternal and fetal assessments, preventive interventions, and person-centred counselling to ensure a positive pregnancy experience [4]. However, accumulating evidence from low- and middle-income countries suggests that high ANC coverage in terms of visit counts does not necessarily translate into high-quality care. Globally, although approximately 88% of pregnant women receive at least one ANC visit, deep disparities persist; many women attend ANC but do not receive key clinical, preventive, or counselling interventions recommended by WHO [5]. Consequently, international tracking agencies shifted their monitoring from a purely volume-based metric to an effective coverage approach. This strategy assesses ANC quality based on the completion of standard clinical interventions rather than contact alone, utilizing composite indices derived from household surveys like the Demographic and Health Surveys (DHS) and Multiple Indicator Cluster Surveys (MICS) [4].

Recent analyses from sub-Saharan Africa show that, although many women attend ANC at least once, effective coverage of quality ANC remains suboptimal. A recent multi-country study using DHS data from 13 sub-Saharan African countries spanning from 2015 to 2021 constructed a composite ANC quality measure based on receipt of 4 essential services including blood pressure measurement, urine testing, blood testing, and iron supplementation, and reported that only 53.8% of women, on average, received all 4 interventions, with national estimates ranging from 11% in Burundi to 82.3% in Cameroon [1]. Both Ameyaw et al. (2024a) and Ayele et al. (2025a) noted that secondary or higher education, marital status, and completing at least four visits significantly increased the odds of receiving quality care[1, 6]. Complementary work that examined clinical process quality using direct observation of ANC consultations in 13 low- and middle-income countries found that providers, on average, completed only about 39% of recommended clinical actions of history taking, examinations, tests, and counselling, with wide variation both within and between countries and poor correlation between structural readiness and actual care delivered [7]. Beyond basic socio-demographics, contemporary literature emphasizes that structural health-system determinants including health insurance coverage, geographic distance, and perceived maternal access barriers like out-of-pocket costs deeply dictate whether a woman receives the full continuum of recommended care actions [8].

Zambia has made important strides in expanding access to maternal health services; however, maternal mortality remains a critical public health challenge, with the 2024 Zambia Demographic and Health Survey (ZDHS) reporting an MMR of 187 maternal deaths per 100,000 live births [9]. National surveys indicate high coverage of at least one ANC visit and substantial utilization of four or more visits. However, concerns persist that the content and quality of care are highly uneven, particularly in rural and impoverished regions [10]. Earlier work in Zambia has predominantly focused on ANC utilisation, such as timing of the first visit and number of contacts, or on broader service readiness, rather than directly measuring the comprehensive quality of clinical, preventive, and counseling care received by the patient. To date, only one national assessment has examined ANC quality in Zambia using older data from the 2005 Health Facility Census and the 2007 Zambia Demographic and Health Survey, highlighting stark socioeconomic and geographic inequities that have not been comprehensively re-evaluated with modern tracking metrics [11]. This leaves a critical, unaddressed 17-year data gap regarding how clinical quality has evolved in the modern healthcare landscape, particularly regarding updated maternal data on counseling interventions, tetanus protection, and preventive drug distribution.

Understanding the current level and distribution of ANC quality is essential to guide efforts to reduce preventable maternal and perinatal deaths and ensure equitable progress toward universal health coverage in Zambia. Specifically, there is a lack of nationally representative evidence on how ANC quality varies across sociodemographic, reproductive, and health-system factors in the post-2016 WHO ANC guideline era. Using data from the recent 2024 Zambia Demographic and Health Survey, this study aims to construct a composite ANC quality index reflecting WHO-recommended clinical, preventive, counseling, and utilization components, and to identify the key factors associated with ANC quality among women with recent live births in Zambia.

## Methods

### Study design and sample

This study used secondary data from the eight round of the Zambia Demographic and Health Survey (ZDHS) which was conducted in 2024. The ZDHS is a nationally representative cross-sectional survey that employed a stratified two-stage cluster sampling design to generate updated estimates of demographic and health indicators especially those tracked by the Sustainable Development Goals (SDGs). Data collection was conducted from 17^th^ January 2024 to 7^th^ July 2024 using 22 teams with each team comprising 12 members: a supervisor, five interviewers, four biomarker technicians and a driver [12]. Data were drawn from the Individual Recode (IR) file of the 2024 Zambia Demographic and Health Survey (ZDHS), which contains one record per eligible woman aged 15–49 years [13]. From a total of 13,951 women interviewed, analysis was restricted to 4,975 women who reported a live birth in the five years preceding the survey and had complete information on antenatal care components received during their most recent pregnancy. Women with missing data on ANC components were excluded from this analysis.

### Variables

The primary outcome was the ANC quality score, a composite index of 15 components representing four domains of WHO-recommended antenatal care: clinical assessment, health counseling and screening, preventive interventions, and ANC utilization [5]. Clinical assessment components included the measurement of weight and blood pressure, urine testing, blood testing, and auscultation of the fetal heartbeat (5 items). Counseling and screening components comprised advice on recommended foods during pregnancy, breastfeeding promotion, guidance on health, diet, and physical activity, screening for vaginal bleeding, and HIV counseling and testing (5 items). Although counseling on danger signs during pregnancy was assessed in the survey, this variable was excluded from the composite index computation due to extensive missing data across the sample. Preventive interventions included iron-containing supplementation, deworming medication, and the receipt of ≥ 2 doses of sulfadoxine-pyrimethamine (IPTp-SP) for malaria prophylaxis (3 items). ANC utilization indicators comprised the attendance of at least four ANC visits and the initiation of ANC within the first trimester (2 items).

Three additional ANC indicators were examined descriptively but excluded from the composite score to avoid duplication or sub-standard indexing. These included the attendance of eight or more ANC visits, the receipt of 2 or more tetanus toxoid doses, and the uptake of only a single dose of IPTp-SP. The latter was excluded because the more comprehensive ≥ 2-dose threshold was already incorporated into the composite score as tracked by the survey timeline.

Each of the 15 ANC intervention components was coded as a binary variable (1 if received, 0 if not received). The continuous ANC quality-of-care score was calculated by summing all 15 components, yielding a range from 0 to 15. A binary outcome, adequate ANC quality, was subsequently created using an 80% threshold (≥12/15 components). This threshold was based on established benchmarks in maternal health quality measurement, where the attainment of approximately four-fifths of recommended interventions is considered indicative of high-quality care [14]. For women with missing data across all 15 components, no score was computed, and these cases were treated as missing.

Independent variables included maternal age, educational attainment, place of residence (urban/rural), province of residence, wealth index, marital status, parity, preceding birth interval, ANC provider type, health insurance status, pregnancy intention (whether the mother wanted the pregnancy then, later, or wanted no more children), and perceived barriers to healthcare access (specifically: distance to the facility, cost of treatment, and obtaining permission to seek care).

### Data management and analysis

“Don’t know” responses were treated as missing prior to analysis. Binary indicators were generated for each of the 15 selected ANC components. Women with missing values across all 15 ANC quality components representing those who did not attend any antenatal care or lacked clinical records were excluded from the final analytic sample, yielding a total sample size of 4,829 women. Data were analyzed using Stata version 19 (StataCorp LLC, College Station, TX, USA). To account for the complex Demographic and Health Survey (DHS) sampling design, primary sampling units, stratification, and probability weights were specified using the svyset command. The individual sampling weight variable (v005) was divided by 1,000,000 to obtain normalized weights, and the svy: prefix was applied to all descriptive, bivariate, and regression analyses to ensure correct variance estimation.

Weighted proportions and 95% confidence intervals (CIs) were calculated to summarize participant characteristics and individual ANC quality indicators. Bivariate associations between participant characteristics and the binary category of ANC adequacy (≥ 80% vs. <80% components) were assessed using survey-adjusted Rao-Scott Pearson chi-square tests. Survey-weighted Poisson regression models were fitted to estimate both crude incidence rate ratios (IRRs) and adjusted incidence rate ratios (aIRRs) with their corresponding 95% CIs for factors associated with the sum of ANC components received as a discrete measure. Unadjusted models were initially calculated for each independent variable, followed by a fully adjusted multivariable model with all pre-specified covariates included simultaneously.

To evaluate the robustness of our primary findings to alternative specifications of ANC quality, sensitivity analyses were conducted utilizing three distinct binary adequacy thresholds: Model 1 (≥ 60%, or ≥ 9/15 components), Model 2 (≥80%, or ≥12/15 components; primary threshold), and Model 3 (≥ 90%, or ≥ 13/15 components). Survey-weighted logistic regression models (svy: logit) were utilized for these sensitivity checks, with results expressed as adjusted odds ratios (aORs) and 95% CIs. Statistical significance across all multivariable models was defined at a 2-tailed alpha level of p < 0.05.

## Results

### Demographic characteristics of the participants

Overall, the analysis included a survey-weighted sample of 4,829 women aged 15 to 49 years. The single largest age group was women aged 20–24 years (25.6%), followed by those aged 25–29 years (23.1%) and adolescents aged 15–19 years (13.3%). Women aged 45–49 years represented less than 1% of the participants (Table 1). The majority of the women had attained a primary education (45.1%), followed closely by a secondary education (41.9%), while the fewest had attained a higher education (5.6%). Structurally, 59.9% of the participants were rural residents, while 40.1% resided in urban areas.

**Table 1.**
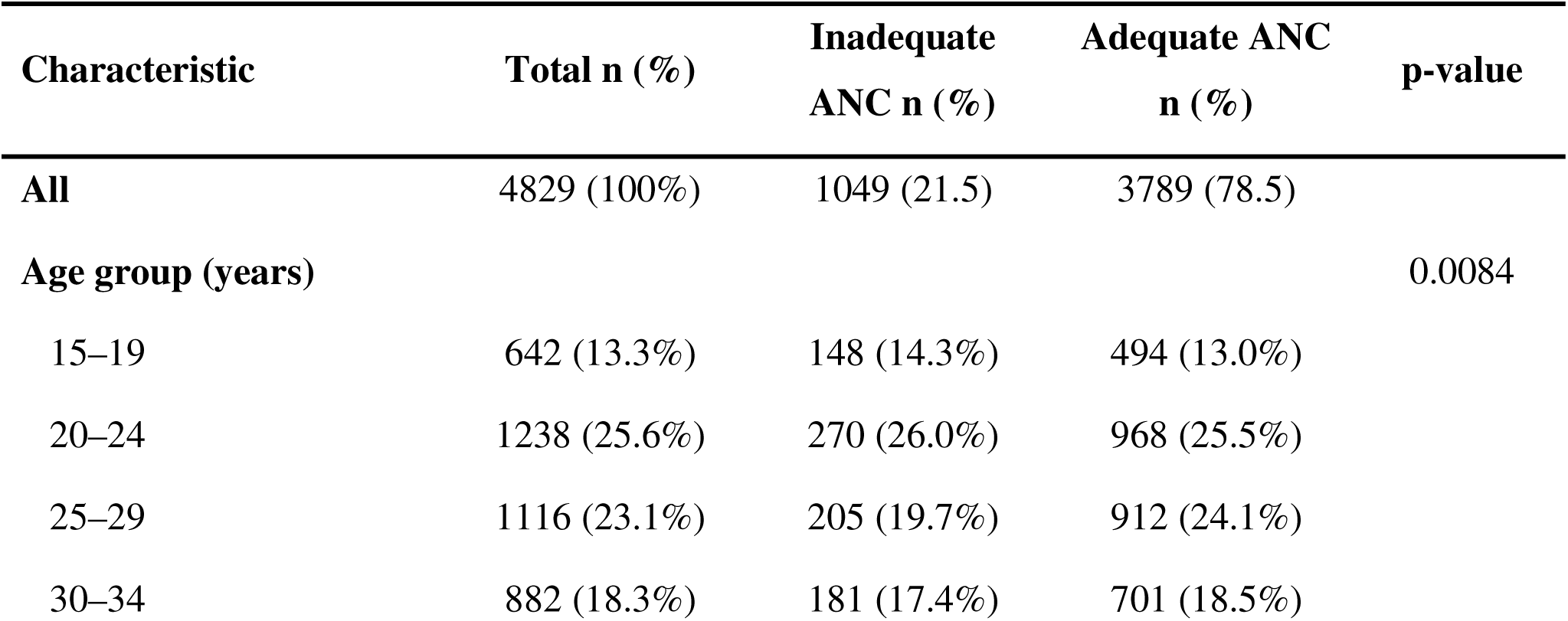
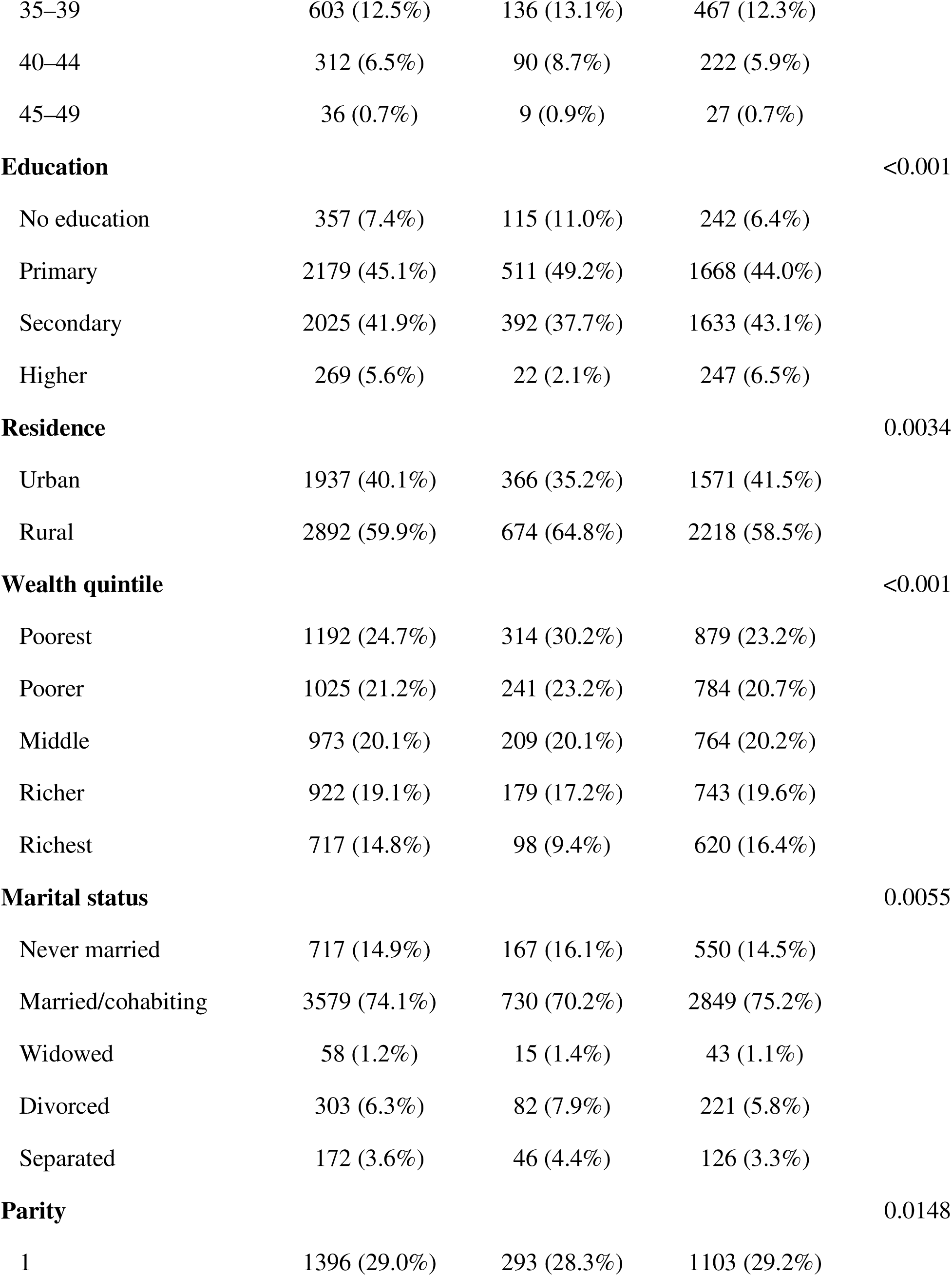
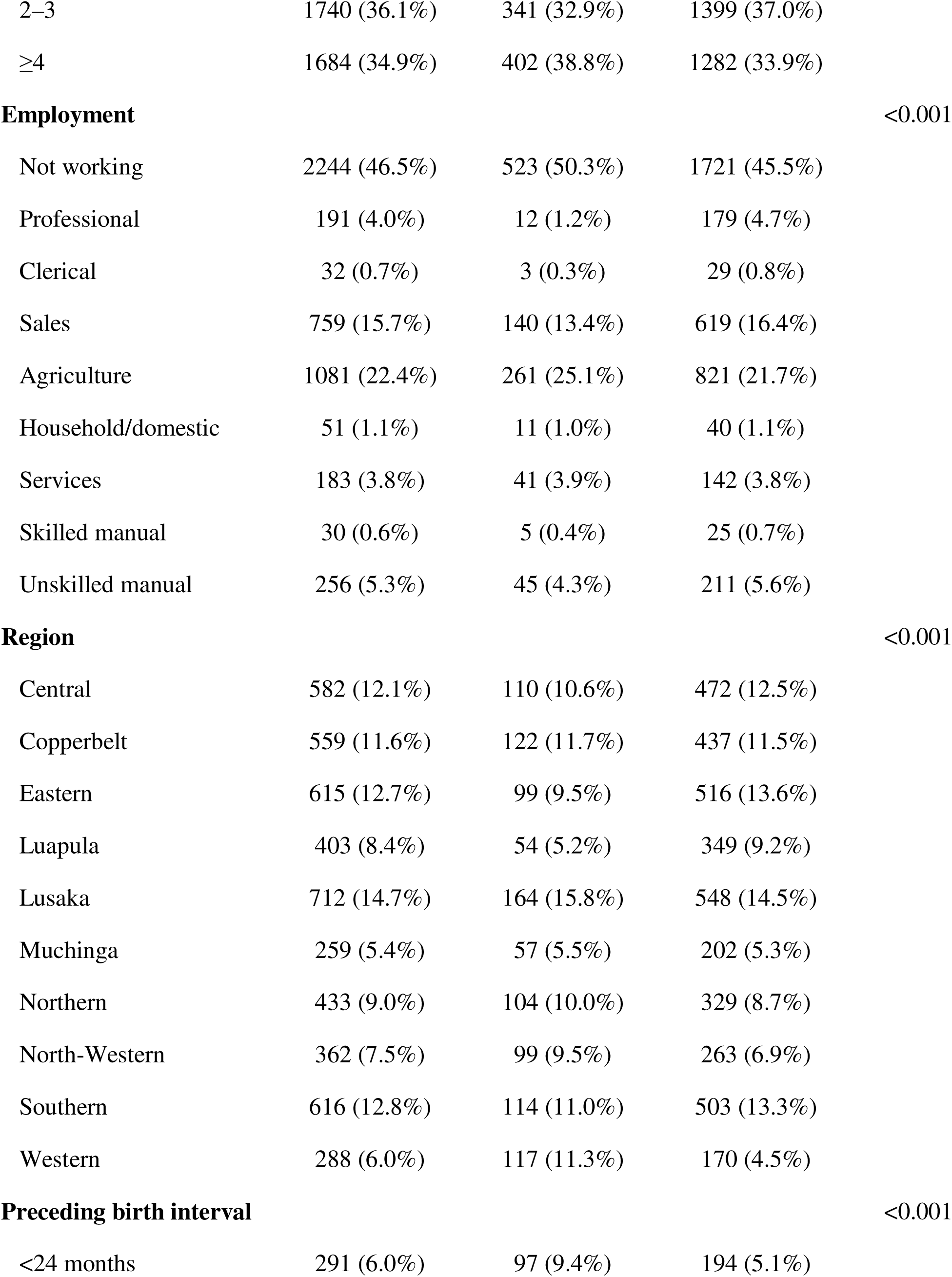
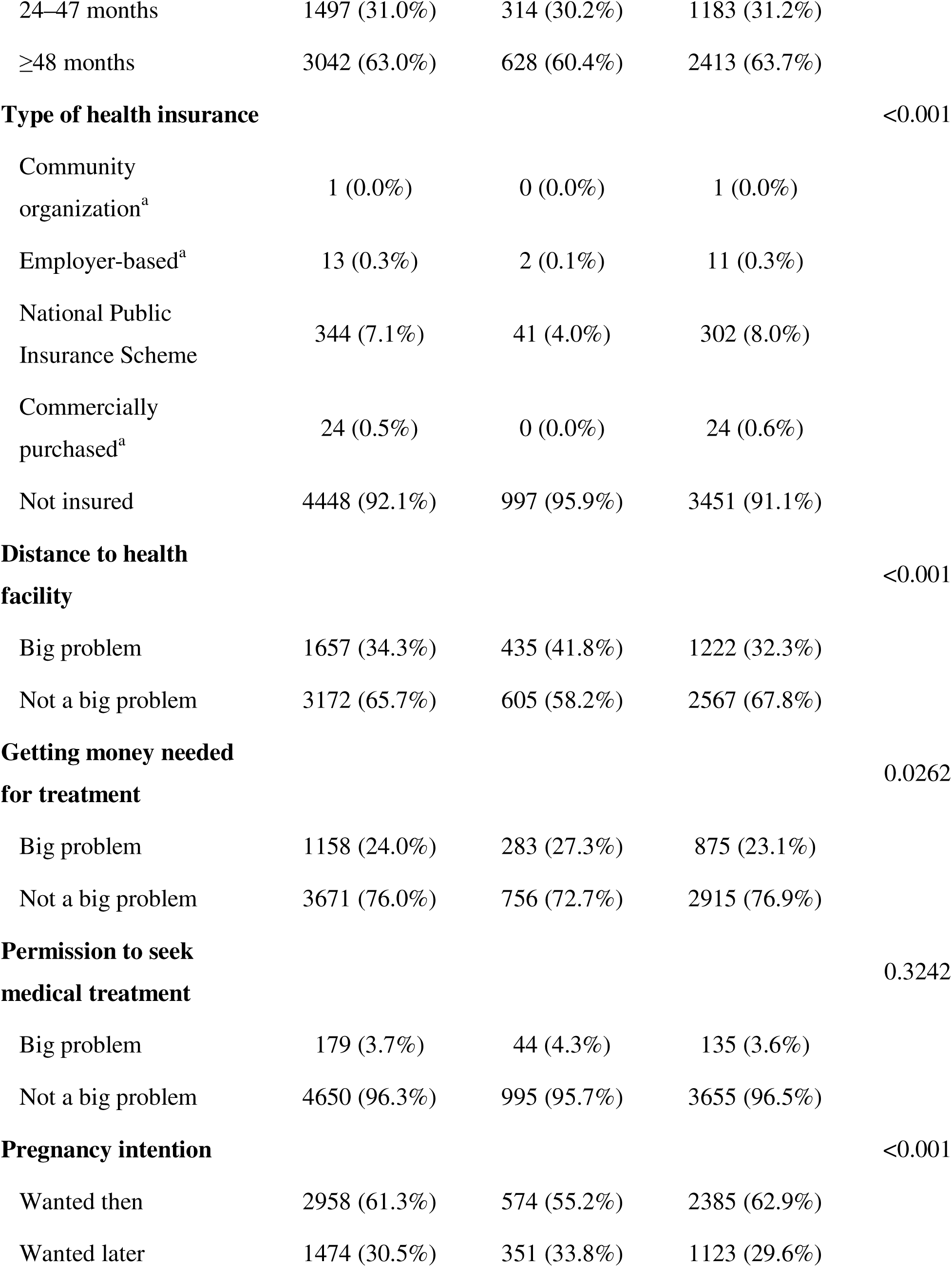
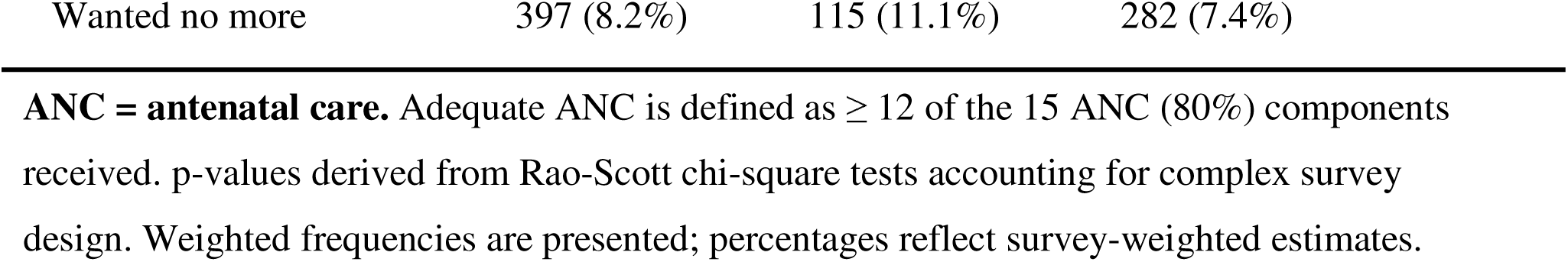
Sociodemographic and maternal characteristics of study participants and distribution of antenatal care quality, Zambia Demographic and Health Survey 2024.

In terms of household wealth quintiles, the poorest quintile comprised the largest proportion of participants (24.7%), followed by the poorer quintile (21.2%), while the richest quintile contained the least (14.8%) (Figure S1). Formally married or cohabiting women formed the overwhelming majority of the sample (74.1%), followed by those who had never married (14.9%), while widowed individuals were the least represented (1.2%). Nearly half of the participants reported not working (46.5%), while the remainder were distributed across various occupations. The largest employed group was engaged in agriculture (22.4%), followed by sales (15.7%), whereas clerical work (0.7%) and skilled manual labor (0.6%) were the least common.

### Quality of antenatal care

Overall, national coverage of quality antenatal care in Zambia was high (Figure1), with 78.5% (95% CI: 77.0–80.0) of women receiving adequate ANC packages, while 21.5% experienced inadequate care as defined by receiving ≤11 of the 15 components. Table 2 presents the weighted baseline estimates for all ANC quality indicators. With respect to ANC utilization, 82.6% (95% CI: 81.3–83.8) of women attended 4 or more ANC visits; however, adherence to the updated WHO recommendation of 8 or more contacts was markedly lower at just 4.8% (95% CI: 4.1–5.6). In addition, fewer than half of the women (47.2%; 95% CI: 45.3–49.0) successfully initiated their first ANC visit within the first trimester.

**Table 2.** Weighted frequency of antenatal care quality indicators among women with a live birth in the past five years, Zambia 2024 DHS. *N = 4812 women (weighted)*

| ANC quality indicator | n (weighted) | % (95% CI) |
| --- | --- | --- |
| **A. ANC utilization** |  |  |
| $\geq 4$ ANC visits | 3975 | 82.6 (81.3-83.8) |
| $\geq 8$ ANC visits (WHO 2016 recommendation) | 231 | 4.8 (4.1-5.6) |
| First ANC visit in first trimester ($\leq 12$ weeks) | 2245 | 47.2 (45.3-49.0) |
| **B. Clinical assessments** |  |  |
| Weight measured | 4648 | 97.7 (97.2-98.1) |
| Blood pressure taken | 4649 | 97.6 (97.0-98.1) |
| Blood sample taken | 4,461 | 93.7 (92.9-94.5) |
| Urine sample taken | 3794 | 79.7 (78.1-81.3) |
| Baby's heart listened to | 4671 | 98.1 (97.7-98.5) |
| **C. Preventive interventions** |  |  |
| Tetanus protection at birth | 3464 | 72.8 (71.2-74.4) |
| Tetanus toxoid ($\geq 2$ doses) | 1176 | 24.9 (23.4-26.4) |
| Iron/folate tablets given or purchased | 4,509 | 93.6 (92.5–94.7) |
| During pregnancy took SP/Fansidar for malaria | 4,536 | 94.2 (93.4–94.9) |
| IPTp-SP received ($\geq 2$ doses) a | 3332 | 73.5 (71.8–75.0) |
| Deworming tablets given | 3432 | 71.6 (69.9–73.2) |
| **D. Counselling and screening** |  |  |
| Talked about health, diet and physical activity | 4,399 | 92.4 (91.4–93.3) |
| Talked about the foods to eat | 4,457 | 93.7 (92.7–94.5) |
| Asked about vaginal bleeding | 4,328 | 90.9 (90.0–91.8) |
| Talked about breastfeeding | 4,380 | 92.2 (91.2–93.0) |
| HIV counselled and tested at ANC | 3114 | 91.9 (90.7–92.9) |
| **E. ANC quality index (composite)** |  |  |
| *Mean quality index score (out of 15) b* | - | 12.5 (12.4–12.6) |
| Adequate quality ($\geq 80\%$ components received) | 3791 | 78.5 (77.0–80.0) |

ANC, antenatal care; CI, confidence interval; IPTp-SP, intermittent preventive treatment in pregnancy with sulfadoxine-pyrimethamine; DHS, Demographic and Health Survey.
All estimates are weighted to account for the complex survey design (PSU, stratification, sample weights).
a While the WHO ideally recommends $\geq 3$ doses of IPTp-SP in sub-Saharan Africa, a threshold of $\geq 2$ doses was utilized for the composite index to align with standard health survey tracking constraints and ensure complete case comparison across the sample timeframe.
b Each indicator scored 1 if received, 0 if not; index = sum of all components (range 0–15).

Individual performance for clinical assessments was exceptionally high: weight measurement, blood pressure recording, and fetal heart auscultation each exceeded 97%, while blood and urine samples were obtained from 93.7% and 79.7% of women, respectively. The coverage of preventive interventions was high for iron/folate supplementation (93.6%) and basic antimalarial prophylaxis (94.2%), although the receipt of ≥ 2 doses of IPTp-SP was lower at 73.5% (95% CI: 71.8–75.0). Approximately 7 out of 10 women received routine deworming tablets (71.6%; 95% CI: 69.9–73.2) and possessed verified tetanus protection at birth (72.8%; 95% CI: 71.2–74.4). Counseling and screening indicators were consistently strong; over 90% of women reported routine clinical discussions regarding maternal diet, breastfeeding, and vaginal bleeding screening, and 91.9% received definitive HIV counseling and testing during their visits (Figure 2).

**Figure 1:**
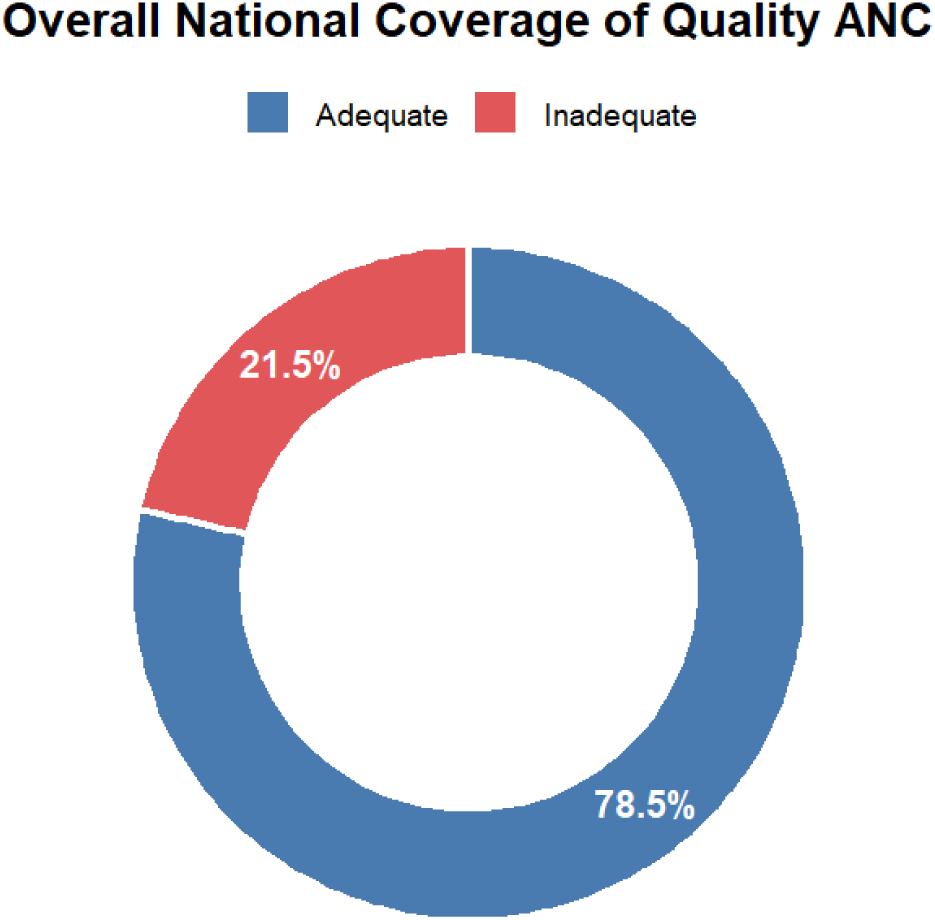
Overall national weighted coverage of Quality ANC

**Figure 2:**
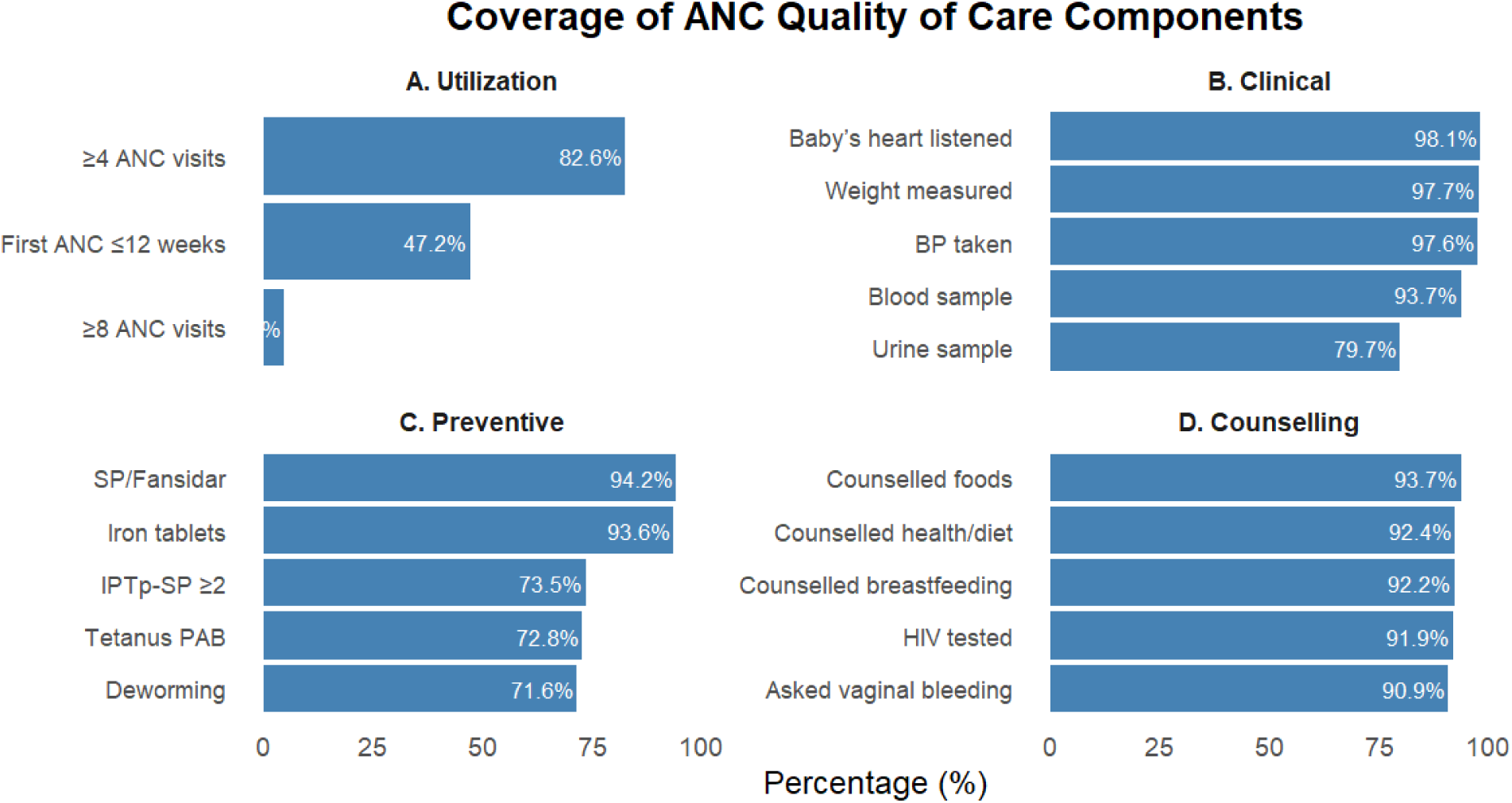
Coverage of ANC Quality of care components

### Bivariate analysis

Bivariate analysis revealed significant associations between care adequacy and several sociodemographic characteristics (Table 1). Educational attainment, wealth quintile, maternal employment status, pregnancy intention, preceding birth interval, health insurance type, and regional variation across all ten provinces (*p* < 0.001) were strongly associated with ANC quality. Place of residence (*p* = 0.003), marital status (*p* = 0.006), age group (*p* = 0.008), parity (*p* = 0.015), and healthcare access barriers such as perceived distance to a facility (*p* < 0.001) and difficulty obtaining money for treatment (*p* = 0.026) were also significantly associated with care outcomes. Conversely, needing permission to seek medical treatment was the only characteristic that failed to show a statistically significant association with ANC quality (*p* = 0.324).

### Multivariable determinants of antenatal care quality

In the multivariable Poisson regression model (Table 3), maternal education remained independently associated with ANC quality after adjusting for all covariates, demonstrating a distinct dose-response pattern across all attainment levels compared to no education (all *p* < 0.001). The significant crude associations observed for household wealth and rural residence were fully attenuated after multivariable adjustment.

**Table 3:**
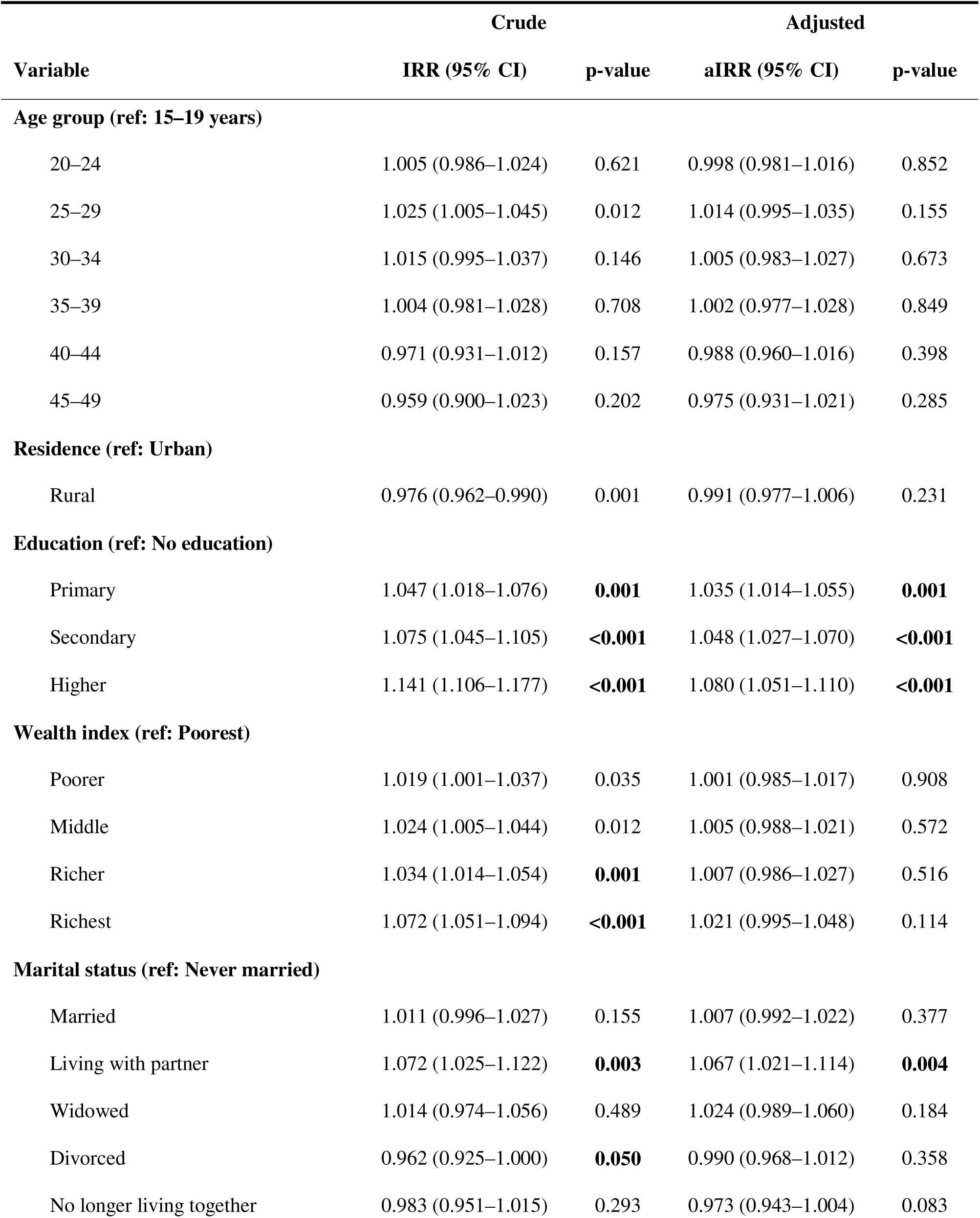
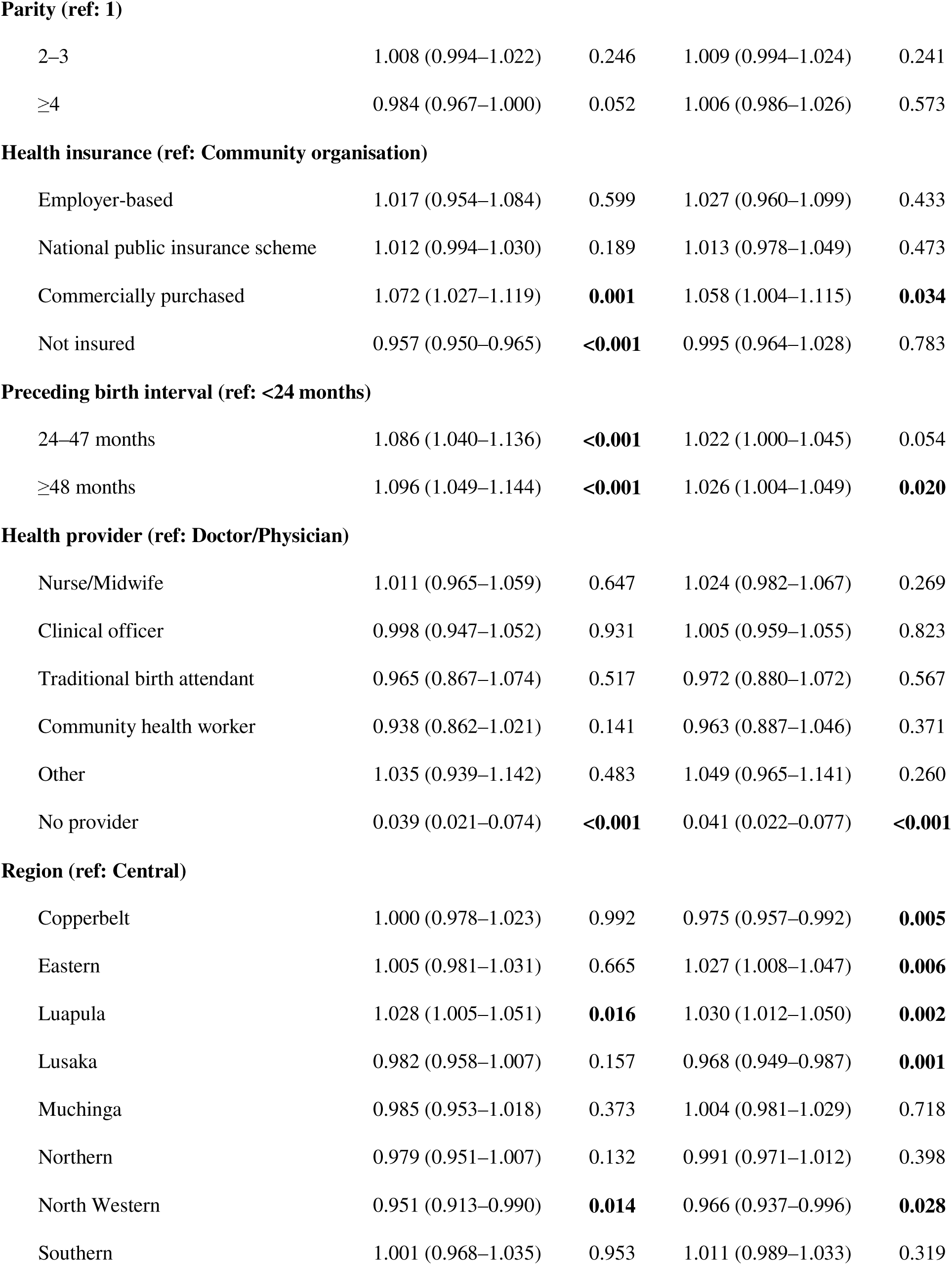
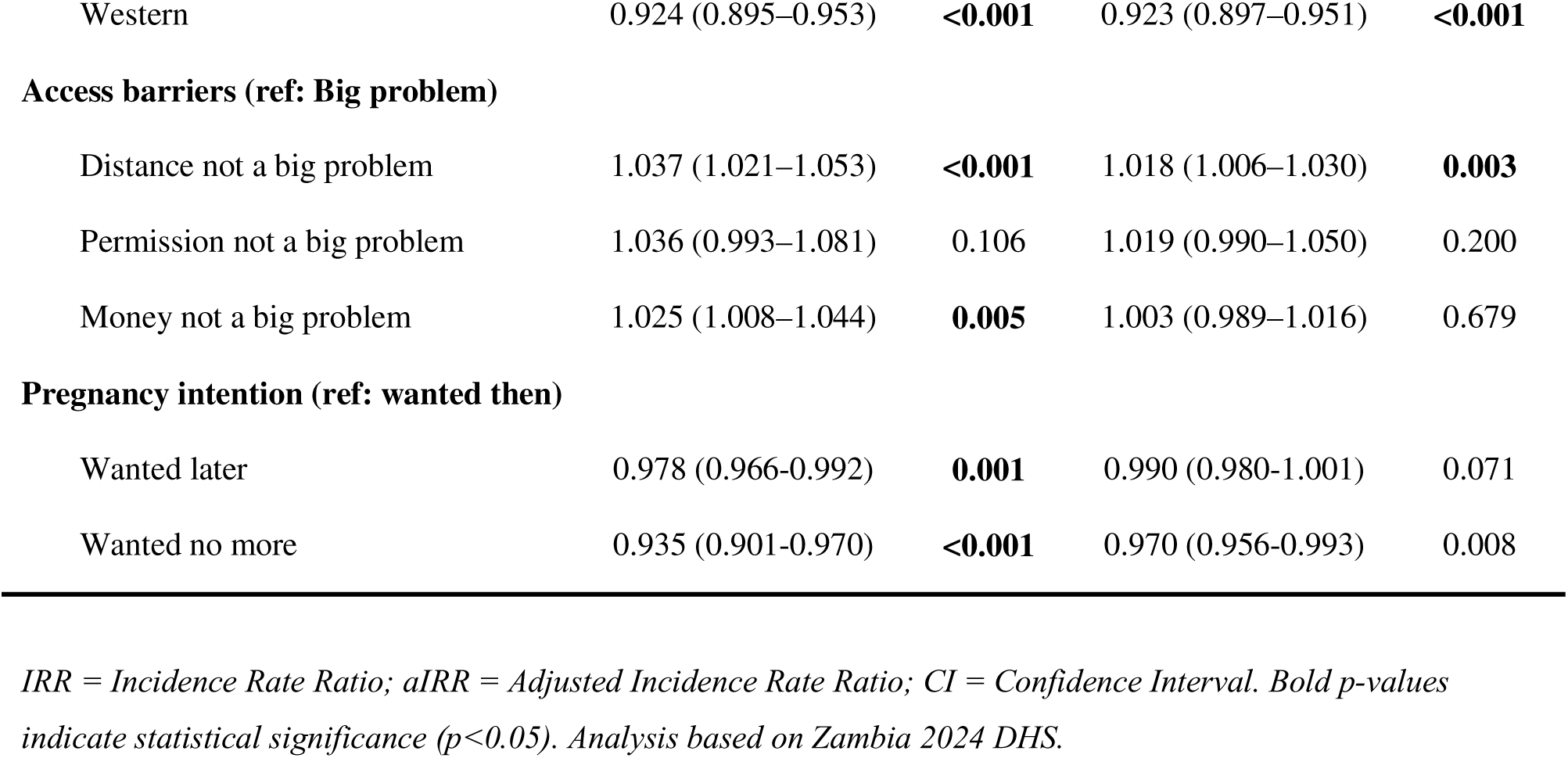
Poisson regression analysis of the determinants of ANC Quality of Care.

| Variable | Crude |  | Adjusted |  |
| --- | --- | --- | --- | --- |
|  | IRR (95% CI) | p-value | aIRR (95% CI) | p-value |
| **Age group (ref: 15–19 years)** |  |  |  |  |
| 20–24 | 1.005 (0.986–1.024) | 0.621 | 0.998 (0.981–1.016) | 0.852 |
| 25–29 | 1.025 (1.005–1.045) | 0.012 | 1.014 (0.995–1.035) | 0.155 |
| 30–34 | 1.015 (0.995–1.037) | 0.146 | 1.005 (0.983–1.027) | 0.673 |
| 35–39 | 1.004 (0.981–1.028) | 0.708 | 1.002 (0.977–1.028) | 0.849 |
| 40–44 | 0.971 (0.931–1.012) | 0.157 | 0.988 (0.960–1.016) | 0.398 |
| 45–49 | 0.959 (0.900–1.023) | 0.202 | 0.975 (0.931–1.021) | 0.285 |
| **Residence (ref: Urban)** |  |  |  |  |
| Rural | 0.976 (0.962–0.990) | 0.001 | 0.991 (0.977–1.006) | 0.231 |
| **Education (ref: No education)** |  |  |  |  |
| Primary | 1.047 (1.018–1.076) | **0.001** | 1.035 (1.014–1.055) | **0.001** |
| Secondary | 1.075 (1.045–1.105) | **<0.001** | 1.048 (1.027–1.070) | **<0.001** |
| Higher | 1.141 (1.106–1.177) | **<0.001** | 1.080 (1.051–1.110) | **<0.001** |
| **Wealth index (ref: Poorest)** |  |  |  |  |
| Poorer | 1.019 (1.001–1.037) | 0.035 | 1.001 (0.985–1.017) | 0.908 |
| Middle | 1.024 (1.005–1.044) | 0.012 | 1.005 (0.988–1.021) | 0.572 |
| Richer | 1.034 (1.014–1.054) | **0.001** | 1.007 (0.986–1.027) | 0.516 |
| Richest | 1.072 (1.051–1.094) | **<0.001** | 1.021 (0.995–1.048) | 0.114 |
| **Marital status (ref: Never married)** |  |  |  |  |
| Married | 1.011 (0.996–1.027) | 0.155 | 1.007 (0.992–1.022) | 0.377 |
| Living with partner | 1.072 (1.025–1.122) | **0.003** | 1.067 (1.021–1.114) | **0.004** |
| Widowed | 1.014 (0.974–1.056) | 0.489 | 1.024 (0.989–1.060) | 0.184 |
| Divorced | 0.962 (0.925–1.000) | **0.050** | 0.990 (0.968–1.012) | 0.358 |
| No longer living together | 0.983 (0.951–1.015) | 0.293 | 0.973 (0.943–1.004) | 0.083 |
| **Parity (ref: 1)** |  |  |  |  |
| 2–3 | 1.008 (0.994–1.022) | 0.246 | 1.009 (0.994–1.024) | 0.241 |
| ≥4 | 0.984 (0.967–1.000) | 0.052 | 1.006 (0.986–1.026) | 0.573 |
| **Health insurance (ref: Community organisation)** |  |  |  |  |
| Employer-based | 1.017 (0.954–1.084) | 0.599 | 1.027 (0.960–1.099) | 0.433 |
| National public insurance scheme | 1.012 (0.994–1.030) | 0.189 | 1.013 (0.978–1.049) | 0.473 |
| Commercially purchased | 1.072 (1.027–1.119) | **0.001** | 1.058 (1.004–1.115) | **0.034** |
| Not insured | 0.957 (0.950–0.965) | **<0.001** | 0.995 (0.964–1.028) | 0.783 |
| **Preceding birth interval (ref: <24 months)** |  |  |  |  |
| 24–47 months | 1.086 (1.040–1.136) | **<0.001** | 1.022 (1.000–1.045) | 0.054 |
| ≥48 months | 1.096 (1.049–1.144) | **<0.001** | 1.026 (1.004–1.049) | **0.020** |
| **Health provider (ref: Doctor/Physician)** |  |  |  |  |
| Nurse/Midwife | 1.011 (0.965–1.059) | 0.647 | 1.024 (0.982–1.067) | 0.269 |
| Clinical officer | 0.998 (0.947–1.052) | 0.931 | 1.005 (0.959–1.055) | 0.823 |
| Traditional birth attendant | 0.965 (0.867–1.074) | 0.517 | 0.972 (0.880–1.072) | 0.567 |
| Community health worker | 0.938 (0.862–1.021) | 0.141 | 0.963 (0.887–1.046) | 0.371 |
| Other | 1.035 (0.939–1.142) | 0.483 | 1.049 (0.965–1.141) | 0.260 |
| No provider | 0.039 (0.021–0.074) | **<0.001** | 0.041 (0.022–0.077) | **<0.001** |
| **Region (ref: Central)** |  |  |  |  |
| Copperbelt | 1.000 (0.978–1.023) | 0.992 | 0.975 (0.957–0.992) | **0.005** |
| Eastern | 1.005 (0.981–1.031) | 0.665 | 1.027 (1.008–1.047) | **0.006** |
| Luapula | 1.028 (1.005–1.051) | **0.016** | 1.030 (1.012–1.050) | **0.002** |
| Lusaka | 0.982 (0.958–1.007) | 0.157 | 0.968 (0.949–0.987) | **0.001** |
| Muchinga | 0.985 (0.953–1.018) | 0.373 | 1.004 (0.981–1.029) | 0.718 |
| Northern | 0.979 (0.951–1.007) | 0.132 | 0.991 (0.971–1.012) | 0.398 |
| North Western | 0.951 (0.913–0.990) | **0.014** | 0.966 (0.937–0.996) | **0.028** |
| Southern | 1.001 (0.968–1.035) | 0.953 | 1.011 (0.989–1.033) | 0.319 |
| Western | 0.924 (0.895–0.953) | <**0.001** | 0.923 (0.897–0.951) | <**0.001** |
| **Access barriers (ref: Big problem)** |  |  |  |  |
| Distance not a big problem | 1.037 (1.021–1.053) | <**0.001** | 1.018 (1.006–1.030) | **0.003** |
| Permission not a big problem | 1.036 (0.993–1.081) | 0.106 | 1.019 (0.990–1.050) | 0.200 |
| Money not a big problem | 1.025 (1.008–1.044) | **0.005** | 1.003 (0.989–1.016) | 0.679 |
| **Pregnancy intention (ref: wanted then)** |  |  |  |  |
| Wanted later | 0.978 (0.966–0.992) | **0.001** | 0.990 (0.980–1.001) | 0.071 |
| Wanted no more | 0.935 (0.901–0.970) | <**0.001** | 0.970 (0.956–0.993) | 0.008 |

*IRR = Incidence Rate Ratio; aIRR = Adjusted Incidence Rate Ratio; CI = Confidence Interval. Bold p-values indicate statistical significance ( $p < 0.05$ ). Analysis based on Zambia 2024 DHS.*

Relationship status emerged as a powerful predictor of care metrics. Women living with a partner received a 6.7% higher absolute count of ANC interventions compared to never-married women (aIRR = 1.067, 95% CI: 1.021–1.114, *p* = 0.004). Similarly, a preceding birth interval of 48 months or more was associated with higher quality scores compared to short intervals under 24 months (aIRR = 1.026, 95% CI: 1.004–1.049, *p* = 0.020). Commercially purchased health insurance was associated with a 5.8% higher care quality score (aIRR = 1.058, 95% CI: 1.004–1.115, *p* = 0.034), whereas the crude risk of being entirely uninsured was attenuated after adjustment.

Having no formal antenatal care provider was associated with drastically lower care scores (aIRR = 0.041, 95% CI: 0.022–0.077, *p* < 0.001). Perceived distance to a health facility not being a big problem was associated with a modest but significant 1.8% higher ANC quality score (aIRR = 1.018, 95% CI: 1.006–1.030, *p* = 0.003). Pronounced regional variation persisted after adjustment, with Western and North-Western provinces showing significantly lower care quality, while Eastern and Luapula provinces demonstrated significantly higher scores relative to Central Province (all *p* < 0.05).

### Sensitivity analysis

Findings from the sensitivity analysis across the three composite outcome thresholds ≥ (60%, ≥ 80%, and ≥ 90%) yielded patterns highly consistent with the main Poisson model (Table S1). The strong dose-response relationship between education and care quality was replicated across all three logistical thresholds. Notably, the effect of formal cohabitation was amplified when examining absolute care adequacy, showing a massive five-fold increase in the odds of achieving the primary ≥ 80% adequacy threshold (Model 2 aOR = 5.01, 95% CI: 1.46–17.16) and retaining strong significance at the top-tier ≥ 90% threshold (Model 3 aOR = 2.28, 95% CI: 1.10–4.75).

The protective effect of longer preceding birth intervals persisted through the ≥ 60% and ≥ 80% models but was attenuated at the strict ≥ 90% threshold. Geographic variation remained an entrenched feature across all thresholds; Western Province exhibited persistently depressed odds of achieving care adequacy across all three models, while associations for other provinces proved threshold-dependent. Perceived distance barriers retained statistical significance only within the primary ≥ 80% adequacy framework. Finally, the negative impact of unintended pregnancy persisted, with women who desired no more children showing significantly lower care quality and reduced odds of achieving high adequacy metrics across both the ≥ 80% and ≥ 90% sensitivity thresholds.

## Discussion

This study examined the quality of ANC received by women aged 15-49 years with a recent live birth in Zambia using a nationally representative sample from the 2024 ZDHS. The analysis applied a composite score of 15 ANC intervention components based on the 2016 WHO guidelines. Our findings demonstrate high overall quality of ANC, as well as high coverage of individual ANC components with over three quarters (78.5%) of women receiving adequate quality ANC and mean score of 12.5. However, attendance of 8 or more ANC visits in line with the national guidelines was strikingly low at 4.8% and less than half of the women-initiated ANC within the first trimester. Education attainment was the strongest predictor of ANC quality and exhibited a dose-response relationship which was consistent across all sensitivity analyses.

Wealth quintile and rural residence were significant in the crude analysis, but the association was attenuated in the adjusted analysis. Additional predictors of higher ANC quality included living with a partner, longer preceding birth interval, commercially purchased health insurance, and absence of distance barriers to care, while unwanted pregnancy and lack of a formal ANC provider were associated with lower quality. Significant regional variation persisted after full adjustment, with Western and North-Western provinces showing consistently lower ANC quality.

Our findings of high ANC quality in Zambia indicate a significant improvement over those from a similar study (also in Zambia) published in 2012, based on data from two decades ago (2005 Health Facility Census and 2007 ZDHS). That study found that only 3% of 45 health facilities met the criteria for optimal ANC service, only 29% of women received good-quality service, and only 8% of mothers attended ANC in the first trimester. This trajectory of improvement is documented in many other sub-Saharan African countries [15], due to the scale-up of skilled birth attendance programmes in Africa [16, 17]. In Zambia specifically, multifaceted interventions have been implemented which include the adoption and integration of the 2016 WHO antenatal care guidelines and their integration into national guidelines and reporting systems (HMIS), adoption and implementation of digital checklists, provision of ANC cards to pregnant women, strengthening mentorships, and development, implementation of Quality of Care Standards, and strengthening referral mechanisms for specialized care [18–20]. Similar improvements in quality of ANC have been documented in other African countries such as Kenya, Benin and Cameroon [15, 21]. Despite gains, critical gaps remain in the frequency of ANC visits and first trimester initiation of ANC indicating that improvements in the content of ANC has not been matched by similar improvements in the number of ANC contacts. Some of the barriers for the inadequate frequency of ANC contacts include limited knowledge of the recommended ANC contacts, high cost of attending the recommended visits, belonging to religious groups the discourage ANC, bad attitude of health workers, and poor quality services characterized by long waiting times and stock-out of medications [22].

Educational attainment emerged as the strongest and most consistent predictor of ANC quality in this study, with a clear dose-response gradient across all levels of schooling relative to women with no formal education. This finding is consistent with a growing body of DHS-based evidence from SSA [15, 23, 24]. Studies examining quality ANC across 13 SSA countries have similarly found that women with secondary or higher education had higher odds of receiving quality ANC compared with those without formal education [15]. Similarly, a 31-country multilevel analysis identified secondary education as a significant individual-level predictor of good-quality ANC services [23]. Education enhances health literacy, thereby increasing their understanding of the importance of ANC, increased health care seeking behaviour and strengthens their capacity to demand comprehensive care during ANC encounters [15]. Educating girls therefore may have a strong ripple effect with downstream consequences for quality maternal health services. For example a cross-sectional study examining female education and maternal health care utilization found that higher maternal education was associated with higher maternal health care utilization which included antenatal utilization, capacity to recognize complications of pregnancy, and fertility practices all of which directly influence quality of maternal health services [25].

Our findings indicate that women on privately purchased commercial insurance had a 5.8% higher ANC quality scores compared to those on community based insurance, a finding that persisted after adjustment for covariates. Although overall health insurance coverage in Zambia remains low at 7.9%, this finding carries important implications for equity. Existing evidence across SSA has established insurance as a strong facilitator of ANC utilization [26, 27]. One multi-country analysis found women with health insurance were nearly three times more likely to attend four or more ANC visits compared to uninsured women [26], yet evidence on its relationship with the content and quality of ANC received remains limited. The present study contributes to filling this gap, demonstrating that insurance is associated with receipt of a more complete package of care including both ANC utilization and content of care. Notably, employer-based and national public insurance were not significantly associated with ANC quality in the adjusted model. This distinction likely reflects differential facility access. Whereas the National Health Insurance Scheme (NHIS) and employer-based schemes primarily channel members to accredited public facilities, commercially purchased plans typically enable access to private providers; the higher ANC quality scores among commercially insured women may partly reflect this differential access to facilities, though this mechanism warrants investigation. Thus, expanding NHIS enrollment alone may yield limited gains in ANC quality unless accompanied by investments to raise service standards in accredited public facilities.

Among all individual-level predictors, absence of a formal ANC provider was associated with the most dramatic reduction in ANC quality (aIRR 0.041). Notably, among women who attended facility-based care, provider cadre did not independently predict quality. Nurses, midwives, clinical officers, and community health workers delivered ANC of equivalent quality to that of physicians, supporting the effectiveness of task-shifting in Zambia’s ANC system. The finding points to a residual layer of systemic exclusion that national quality improvement interventions have not yet reached.

Unwanted pregnancy among women who wanted no more children was associated with lower ANC quality scores. This is consistent with evidence from SSA showing that mistimed and unwanted pregnancies are strongly associated with late ANC attendance and fewer visits, representing a critical risk to achieving adequate ANC quality [28]. Evidence from a qualitative study conducted in Kenya found that unintended pregnancy leads to poor ANC uptake through reduced partner/family support [29]. Additionally, women with unintended pregnancies may lack the motivation to engage with ANC leading to lower utilization and consequently missed ANC intervention components. These findings underscore the importance of strengthening family planning services across the reproductive health continuum to prevent unintended pregnancies, while ensuring that women who experience unwanted pregnancies receive non-judgmental, supportive ANC to maximize engagement in care.

Significant regional variation in ANC quality persisted in the adjusted model, with Western and North-Western provinces showing consistently lower ANC scores while Eastern and Luapula provinces showed higher ANC scores relative to Central province. This pattern is consistent with prior Zambian evidence, which has demonstrated that women in Western province are less likely to complete the continuum of care for maternity care [30]. The province is further characterized by difficult geographic and climatic conditions that negatively impact household income, leading to poverty and inadequate access to quality antenatal care [30]. Geospatial analyses of ANC utilization in Zambia have also documented notable provincial disparities, with North-Western province among those recording higher proportions of inadequate ANC utilization [31]. The higher ANC quality scores in Eastern and Luapula provinces may in part be attributed to the impact of the Saving Mothers Giving Life (SMGL) programme which targeted districts in both provinces [32]. The SMGL was a multifaceted maternal and newborn health intervention implemented in Uganda and Zambia and reduced maternal mortality by 44% in Uganda and 41 in Zambia respectively through a health systems approach at district and health facility level [32]. The findings underscore the need for differentiated quality improvement strategies that address structural barriers in underperforming provinces.

### Strengths and Limitations

A major strength of this study is its use of a large nationally representative sample from a recent 2024 ZDHS, therefore our findings reflect the current state of maternal healthcare in Zambia. The stability of our regression findings across three sensitivity analysis thresholds confirms the internal validity of our findings. However, we acknowledge the following limitations. First, the ZDHS relies on maternal self-reporting of ANC interventions components, making the data susceptible to recall bias. Second, the cross-sectional nature of the survey design indicates that we cannot definitively establish causal relationships. Third, for diagnostics, the variables indicate whether blood or urine samples were collected, but we were unable to verify whether the full panel of blood and urine tests was performed and whether/how they were used for clinical decision-making. Finally, there was missing data for some of the recommended ANC intervention components, such as counselling on danger signs, birth plan/preparedness, and family planning, these were not included in the composite score.

## Conclusion

This study demonstrates that Zambia has achieved relatively high overall ANC quality, reflecting major gains over the past two decades from national investments in maternal health programmes. However, critical gaps in ANC utilisation (four and eight ANC contacts) and in initiation of ANC in the first trimester reveal that improvements in the content of ANC have outpaced gains in service utilisation. Education emerged as the major driver of quality ANC, highlighting that investment in girls’ education carries downstream effects on maternal health quality-of-care indicators, which in turn affect maternal and newborn outcomes. Our findings on regional disparities in ANC component scores indicate that progress has not been uniform across the country. Achieving universal quality ANC requires the implementation of differentiated quality-improvement interventions that address the unique barriers of the different provinces and consider the integration of family planning with antenatal services.

## Supporting information

Figure S1: Supplementary figure showing coverage of adequate Antenatal Care Quality of Care Components in Zambia

Table S1. Sensitivity analysis showing adjusted odds ratios for adequate antenatal care quality across three composite outcome thresholds

## Data Availability

The dataset analysed during the current study are available through the Demographic and Health Survey program website, www.dhsprogram.com, upon request and following registration and approval by the DHS Program.

https://dhsprogram.com/data/dataset_admin/login_main.cfm?CFID=268439998&CFTOKEN=6d26aff0e72eda0e-1F0670B5-F8EF-7D5F-F8CDBFD905C71379

## List of abbreviations

ANC: Antenatal care
aIRR: Adjusted Incidence Rate Ratio
aOR: Adjusted Odds Ratio
CI: Confidence Interval
DHS: Demographic and Health Survey
IRR: Incidence Rate Ratio
IPTp-SP: Intermittent Preventive Treatment in pregnancy with Sulfadoxine-Pyrimethamine
NHIS: National Health Insurance Scheme
OR: Odds Ratio
SMGL: Saving Mothers Giving Life
SSA: sub-Saharan Africa
WHO: World Health Organization
ZDHS: Zambia Demographic and Health Survey

## Declarations

## Ethics approval and consent to participate

This study was a secondary analysis of publicly available and de-identified data from the 2024 ZDHS. Access to the dataset was granted by the DHS Program after officially requesting the dataset through the DHS program (www.dhsprogram.com). Ethical approval for the original survey and instruments was obtained from the Institutional Review Boards (IRBs) of ICF and the National Health Research and Training Institute and the National Health Research Authority in Zambia. Written informed consent was obtained from all participants prior to data collection in the original survey. As this analysis involved only de-identified secondary data with no contact with participants, no additional ethical approval was required. The dataset is freely available to researchers upon registration and approval through the DHS program website.

## Consent for publication

Not applicable

## Author’s contributions

Both PMT and LN conceptualised the study and developed the research question. PMT conducted the data analysis. Both authors drafted the manuscript and reviewed and approved the final version.

## Acknowledgements

Not applicable

## Supplementary files

### Additional file 1

**Table S1.**
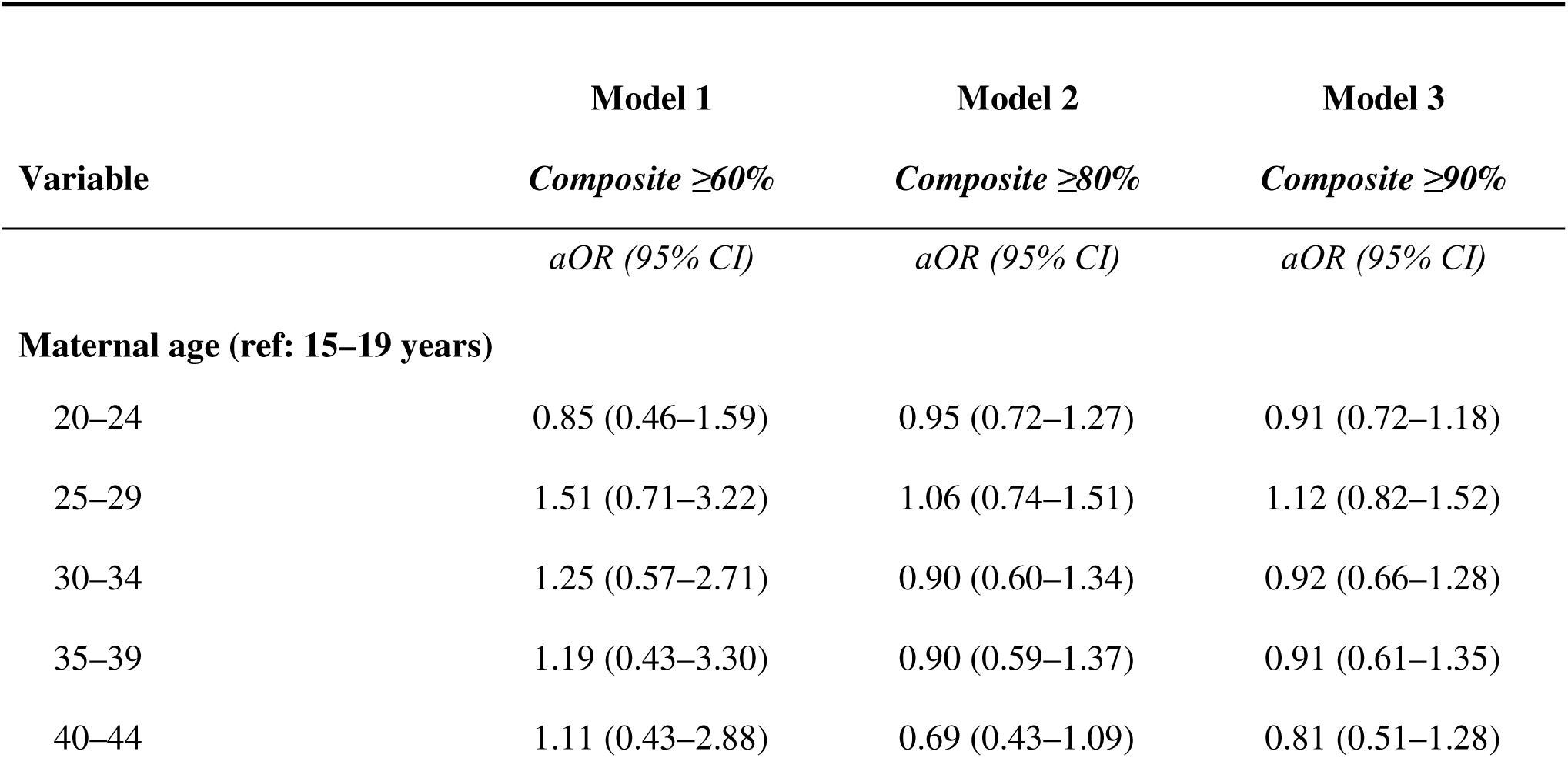
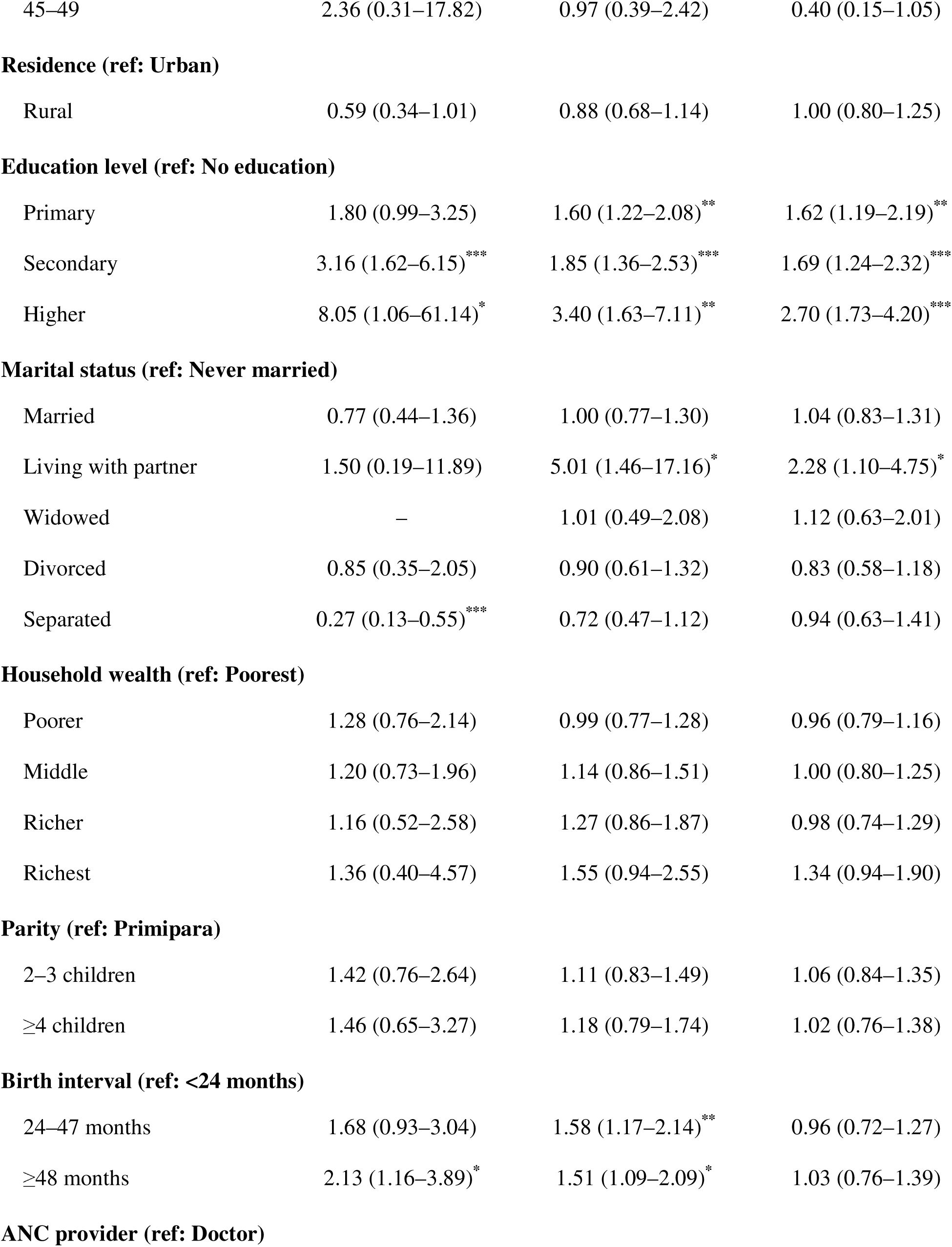
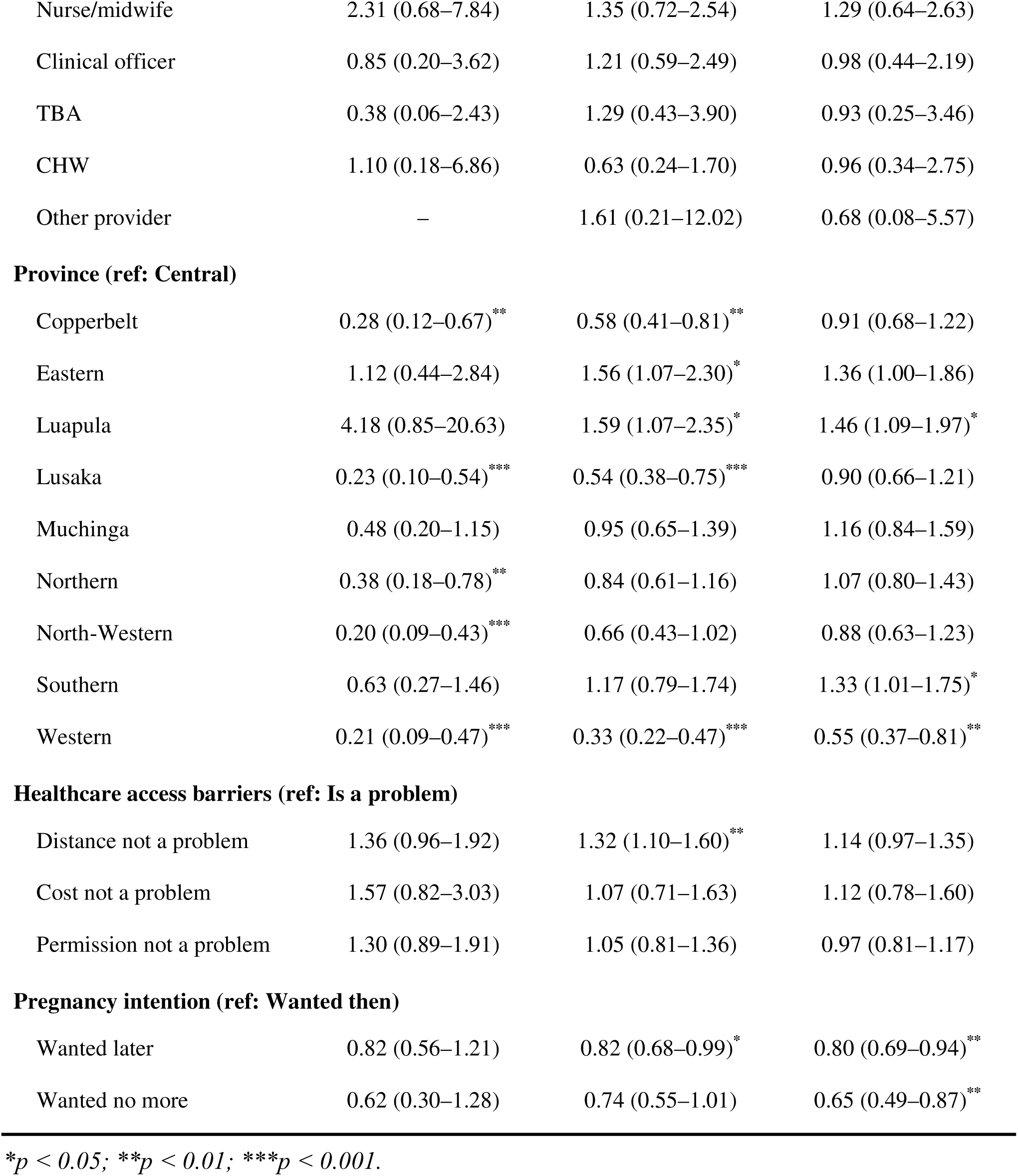
Sensitivity analysis showing adjusted odds ratios for adequate antenatal care quality across three composite outcome thresholds, Zambia Demographic and Health Survey 2024.

**Figure S1:**
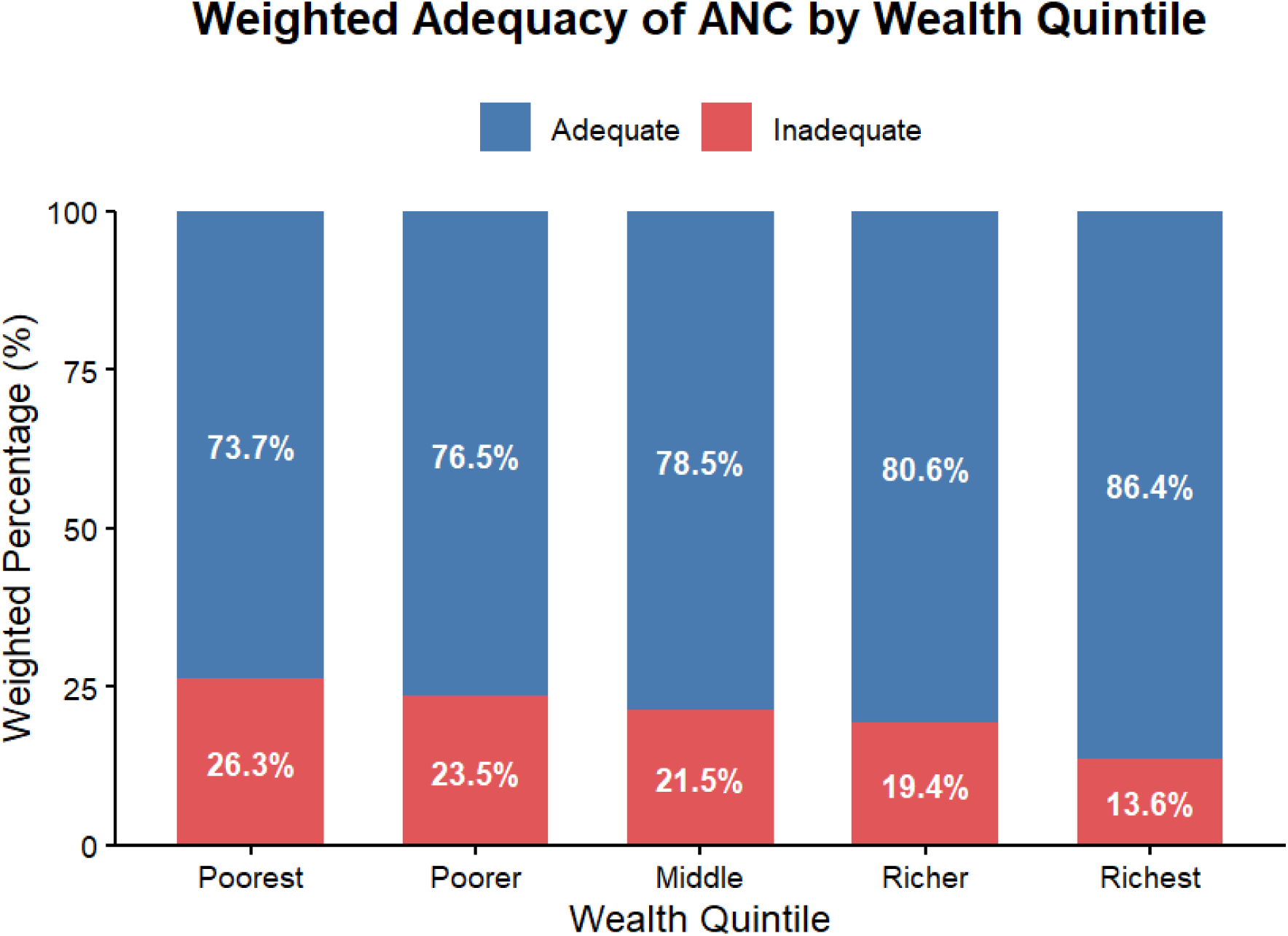
Supplementary figure showing coverage of adequate Antenatal Care Quality of Care Components in Zambia.

