## Supplementary material for "High coverage, persistent gaps: quality of Antenatal Care and its determinants in Zambia based on the 2024 Demographic and Health Survey": Figure S1: Supplementary figure showing coverage of adequate Antenatal Care Quality of Care Components in Zambia

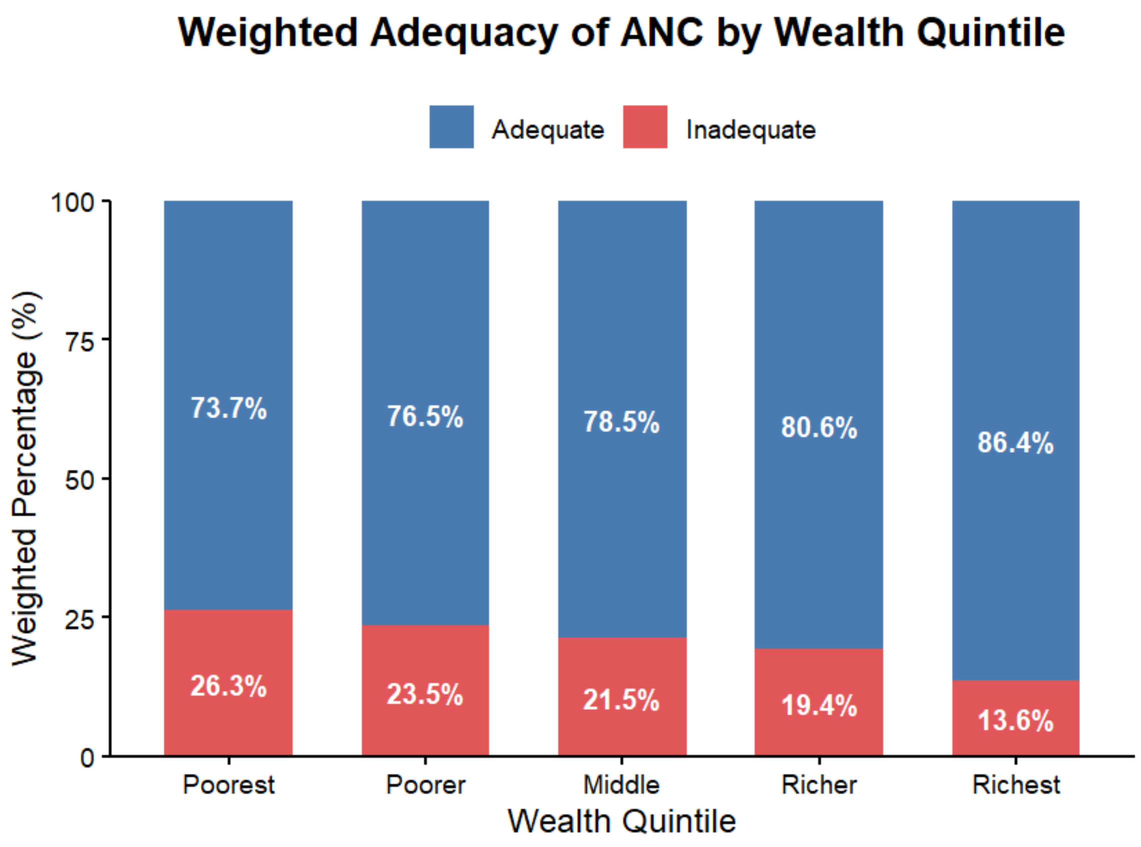


**Figure S1: Supplementary figure showing coverage of adequate Antenatal Care Quality of Care Components in Zambia**
