## Supplementary material for "High coverage, persistent gaps: quality of Antenatal Care and its determinants in Zambia based on the 2024 Demographic and Health Survey": Table S1. Sensitivity analysis showing adjusted odds ratios for adequate antenatal care quality across three composite outcome thresholds

**Supplementary files**

**Table S1.** Sensitivity analysis showing adjusted odds ratios for adequate antenatal care quality across three composite outcome thresholds, Zambia Demographic and Health Survey 2024.

|  | **Model 1** | **Model 2** | **Model 3** |
| --- | --- | --- | --- |
| **Variable** | ***Composite ≥60%*** | ***Composite ≥80%*** | ***Composite ≥90%*** |
|  | *aOR (95% CI)* | *aOR (95% CI)* | *aOR (95% CI)* |
| **Maternal age (ref: 15–19 years)** | | | |
| 20–24 | 0.85 (0.46–1.59) | 0.95 (0.72–1.27) | 0.91 (0.72–1.18) |
| 25–29 | 1.51 (0.71–3.22) | 1.06 (0.74–1.51) | 1.12 (0.82–1.52) |
| 30–34 | 1.25 (0.57–2.71) | 0.90 (0.60–1.34) | 0.92 (0.66–1.28) |
| 35–39 | 1.19 (0.43–3.30) | 0.90 (0.59–1.37) | 0.91 (0.61–1.35) |
| 40–44 | 1.11 (0.43–2.88) | 0.69 (0.43–1.09) | 0.81 (0.51–1.28) |
| 45–49 | 2.36 (0.31–17.82) | 0.97 (0.39–2.42) | 0.40 (0.15–1.05) |
| **Residence (ref: Urban)** | | | |
| Rural | 0.59 (0.34–1.01) | 0.88 (0.68–1.14) | 1.00 (0.80–1.25) |
| **Education level (ref: No education)** | | | |
| Primary | 1.80 (0.99–3.25) | 1.60 (1.22–2.08)**^**^** | 1.62 (1.19–2.19)**^**^** |
| Secondary | 3.16 (1.62–6.15)**^***^** | 1.85 (1.36–2.53)**^***^** | 1.69 (1.24–2.32)**^***^** |
| Higher | 8.05 (1.06–61.14)**^*^** | 3.40 (1.63–7.11)**^**^** | 2.70 (1.73–4.20)**^***^** |
| **Marital status (ref: Never married)** | | | |
| Married | 0.77 (0.44–1.36) | 1.00 (0.77–1.30) | 1.04 (0.83–1.31) |
| Living with partner | 1.50 (0.19–11.89) | 5.01 (1.46–17.16)**^*^** | 2.28 (1.10–4.75)**^*^** |
| Widowed | – | 1.01 (0.49–2.08) | 1.12 (0.63–2.01) |
| Divorced | 0.85 (0.35–2.05) | 0.90 (0.61–1.32) | 0.83 (0.58–1.18) |
| Separated | 0.27 (0.13–0.55)**^***^** | 0.72 (0.47–1.12) | 0.94 (0.63–1.41) |
| **Household wealth (ref: Poorest)** | | | |
| Poorer | 1.28 (0.76–2.14) | 0.99 (0.77–1.28) | 0.96 (0.79–1.16) |
| Middle | 1.20 (0.73–1.96) | 1.14 (0.86–1.51) | 1.00 (0.80–1.25) |
| Richer | 1.16 (0.52–2.58) | 1.27 (0.86–1.87) | 0.98 (0.74–1.29) |
| Richest | 1.36 (0.40–4.57) | 1.55 (0.94–2.55) | 1.34 (0.94–1.90) |
| **Parity (ref: Primipara)** | | | |
| 2–3 children | 1.42 (0.76–2.64) | 1.11 (0.83–1.49) | 1.06 (0.84–1.35) |
| ≥4 children | 1.46 (0.65–3.27) | 1.18 (0.79–1.74) | 1.02 (0.76–1.38) |
| **Birth interval (ref: <24 months)** | | | |
| 24–47 months | 1.68 (0.93–3.04) | 1.58 (1.17–2.14)**^**^** | 0.96 (0.72–1.27) |
| ≥48 months | 2.13 (1.16–3.89)**^*^** | 1.51 (1.09–2.09)**^*^** | 1.03 (0.76–1.39) |
| **ANC provider (ref: Doctor)** | | | |
| Nurse/midwife | 2.31 (0.68–7.84) | 1.35 (0.72–2.54) | 1.29 (0.64–2.63) |
| Clinical officer | 0.85 (0.20–3.62) | 1.21 (0.59–2.49) | 0.98 (0.44–2.19) |
| TBA | 0.38 (0.06–2.43) | 1.29 (0.43–3.90) | 0.93 (0.25–3.46) |
| CHW | 1.10 (0.18–6.86) | 0.63 (0.24–1.70) | 0.96 (0.34–2.75) |
| Other provider | – | 1.61 (0.21–12.02) | 0.68 (0.08–5.57) |
| **Province (ref: Central)** | | | |
| Copperbelt | 0.28 (0.12–0.67)**^**^** | 0.58 (0.41–0.81)**^**^** | 0.91 (0.68–1.22) |
| Eastern | 1.12 (0.44–2.84) | 1.56 (1.07–2.30)**^*^** | 1.36 (1.00–1.86) |
| Luapula | 4.18 (0.85–20.63) | 1.59 (1.07–2.35)**^*^** | 1.46 (1.09–1.97)**^*^** |
| Lusaka | 0.23 (0.10–0.54)**^***^** | 0.54 (0.38–0.75)**^***^** | 0.90 (0.66–1.21) |
| Muchinga | 0.48 (0.20–1.15) | 0.95 (0.65–1.39) | 1.16 (0.84–1.59) |
| Northern | 0.38 (0.18–0.78)**^**^** | 0.84 (0.61–1.16) | 1.07 (0.80–1.43) |
| North-Western | 0.20 (0.09–0.43)**^***^** | 0.66 (0.43–1.02) | 0.88 (0.63–1.23) |
| Southern | 0.63 (0.27–1.46) | 1.17 (0.79–1.74) | 1.33 (1.01–1.75)**^*^** |
| Western | 0.21 (0.09–0.47)**^***^** | 0.33 (0.22–0.47)**^***^** | 0.55 (0.37–0.81)**^**^** |
| **Healthcare access barriers (ref: Is a problem)** | | | |
| Distance not a problem | 1.36 (0.96–1.92) | 1.32 (1.10–1.60)**^**^** | 1.14 (0.97–1.35) |
| Cost not a problem | 1.57 (0.82–3.03) | 1.07 (0.71–1.63) | 1.12 (0.78–1.60) |
| Permission not a problem | 1.30 (0.89–1.91) | 1.05 (0.81–1.36) | 0.97 (0.81–1.17) |
| **Pregnancy intention (ref: Wanted then)** | | | |
| Wanted later | 0.82 (0.56–1.21) | 0.82 (0.68–0.99)**^*^** | 0.80 (0.69–0.94)**^**^** |
| Wanted no more | 0.62 (0.30–1.28) | 0.74 (0.55–1.01) | 0.65 (0.49–0.87)**^**^** |

*aOR, adjusted odds ratio; CI, confidence interval; TBA, traditional birth attendant; CHW, community health worker. Models differ by outcome threshold for the composite ANC quality index: Model 1 = ≥60% (≥9/15 components); Model 2 = ≥80% (≥12/15 components, primary analysis); Model 3 = ≥90% (≥13/15 components). All models fitted using survey-weighted logistic regression (svyglm) with Taylor-series linearisation to account for complex sampling. Reference categories given in parentheses in group headers. – denotes that the estimate could not be computed owing to an empty stratum. *p < 0.05; **p < 0.01; ***p < 0.001.*
